# Disability Documentation in the National Health Interview Survey and Its Consequence: Comparing the American Community Survey to the Washington Group Disability Measures

**DOI:** 10.1101/2023.10.16.23297081

**Authors:** Scott D. Landes, Bonnielin K. Swenor, Nastassia Vaitsiakhovich

## Abstract

**Background and Objective.:** The objective of the National Health Interview Survey (NHIS) is to provide data that can be used to monitor the health of the US population. In this study, we evaluate whether the disability questions currently used in the NHIS – the Washington Group questions – threaten the ability of this survey to fulfil its stated objective for disabled people.

**Methods.:** Data were from the 2011-2012 NHIS with linkage to mortality status through 2019. We examined the percentage and characteristics of people reporting a disability in the American Community Survey (ACS) disability questions who were documented as such in the Washington Group (WG) disability questions. We then examined the consequence of use of the WG, as opposed to the ACS questions, on estimates of disability prevalence and comparative mortality risk.

**Results.:** We find that when compared to their predecessor, the American Community Survey disability questions, the Washington Group questions account for less than half of disabled people, primarily documenting disabled people with a more than one disability status, but excluding many disabled people with only one disability status. As a result of these exclusions, disability prevalence rates based on the Washington Group questions underestimate the size of the disabled population in the US, and overestimate the comparatively higher mortality risk associated with disability status.

**Conclusions.:** These results underscore the need to re-evaluate the disability questions used in the NHIS, and invest in the development of improved and expanded disability questionnaires for use in national surveys.

## Background

The National Health Interview Survey (NHIS) provides annual nationally representative cross-sectional data on health outcomes and healthcare access for the non-institutionalized US population, as well as data on mortality patterns through linkage to the National Death Index (NDI). As such, it is a crucial element of US public health strategy, providing the empirical evidence policymakers need to understand population health and mortality trends and shape effective health policy. The Centers for Disease Control and Prevention (CDC) aptly describes the main objective of the NHIS as providing data that can be used to “monitor the health of the US population.”^1^ In this study we – three disabled scholars – examine whether the questions currently used to measure disability status in the NHIS permit this objective to be fulfilled for disabled people.

Over 67 million (26.8%) adults in the United States are disabled.^2^ A growing amount of evidence reveals that disabled people in the US experience persistent health inequities,^3–6^ inclusive of: higher prevalence of chronic health conditions;^7,8^ structural barriers to accessing healthcare services (e.g., physical and communication barriers, clinician bias);^3,9–12^ higher likelihood of foregone services due to cost;^13,14^ and disability discrimination in medical settings.^15,16^ The false equivalence of disability and adverse health that lingers within the medical system likely informs these ongoing inequities.^3,17^ As a result, the mainstream agenda of public health and medical research is often built around preventing and treating impairments.^18,19^ This narrow perspective overlooks structural ableism, which largely contributes to health inequities.^18^ In this context, it is important to prioritize research on improving health and well-being for disabled people.^17^ However, there is no way to do that without accurate disability data.^19,20^

The current and historic absence of adequate disability data is a product of the social injustice that enshrines health inequities.^19,21^ Closing this data gap necessitates a methodological standard for identifying the disabled population across data collections.^22^ When disability status is not measured accurately, it is difficult to determine whether observed patterns of health inequities reflect existing realities or methodological variations.^22^ Thus, inclusionary evidence-based policies cannot be developed without standardized and accurate measures of disability status within nationally-representative surveys.^19,22,23^

Recognizing the importance of harmonized disability data, Section 4302 of the Affordable Care Act requires establishing a data collection standard for tracking federal information on disability status.^24^ Currently, there are two sets of disability questions used in nationally representative surveys in the US. The disability questions recommended for use by the Department of Health and Human Services (HHS)^24^ are those currently used in the American Community Survey (ACS) and currently or previously used in at least 15 other national surveys administered by federal agencies.^25^ A second set of disability questions developed in 2006 by the Washington Group on Disability Statistics (WG), a City Group chartered by the United Nations Statistical Commission,^26^ are currently used in two national surveys.^26^

The ACS questions were fielded in the NHIS from 2008 to 2019 to children and adults selected to receive the Family Disability test questions, but have not been included in the NHIS since 2019.^27^ The Washington Group-Short Set (WG-SS) questions were introduced to the NHIS in 2010, fielded to Sample Adult subsamples through 2017, then fielded to all Sample Adults from 2018 forward.^27^ Two studies report that the ACS disability questions identify 1.5 to 2 times more disabled adults than the WG-SS questions in the NHIS.^26,28^ However, direct comparison of the performance of the ACS and WG-SS in estimating disability prevalence among adults using NHIS data is incomplete, as these studies either used non-overlapping samples from different years^28^ – meaning one subsample from year X was asked the ACS questions, a separate subsample from year Y the WG-SS questions – or when using the same subsamples from the same year, did not weight data,^26^ a necessary step in calculating estimates with NHIS data.

Despite acknowledged concerns that use of the WG-SS disability questions in the NHIS results in an undercount of disabled people, sufficient examination has not been conducted to determine which disabled people are excluded when using the WG-SS questions, and whether this threatens the ability of the NHIS to achieve its objective. To address these concerns, we pursue three aims in this study. First, we evaluate whether the undercounting of disabled people in the NHIS with the WG-SS disability questions is due to misclassification associated with disability status(es) or certain socioeconomic characteristics. Second, we compare the effect of using the WG-SS questions in the NHIS, as opposed to the ACS questions, on our understanding disability prevalence in the US. Finally, we analyze the effect of using the WG-SS, as opposed to the ACS, on estimates of one key population health outcome,^29^ comparative mortality risk between disabled and nondisabled people.

## Methods

### Data and sample

We used data from the 2011 and 2012 NHIS Sample Adult Files obtained from IPUMS.^27^ These were the only years in which NHIS fielded both the ACS and WG-SS disability questions to a subset of sample adults. After excluding the 594 cases without a response to either the ACS or WG-SS disability questions (N = 594), our final analytic sample included 24,694 cases. Examination of mortality outcomes used the NHIS linked mortality files (NHIS-LMF) for those respondents in the study who had an eligible mortality status through 2019 (N = 24,252).

### Measures

#### Disability status

Both the ACS disability questions and WG-SS are informed by the International Classification of Functioning, Disability, and Health (ICF).^30,31^ The ACS disability measures indicate whether the respondents status in six domains of functioning: being deaf or having serious difficulty hearing (hearing); being blind or having serious difficulty seeing (vision); having serious difficulty concentrating or remembering (concentrating/remembering); having difficulty walking or climbing stairs (mobility); having difficulty dressing or bathing (selfcare); and having difficulty doing errands (Instrumental Activities of Daily Living). ^32^ Each measure is coded dichotomously (yes, no).

The WG-SS disability measures indicate the amount of difficulty the respondent reported having in six core areas.: vision (vision); hearing (hearing); walking or climbing stairs (mobility); remembering or concentrating (concentrating/remembering); washing or dressing (selfcare); and communicating in usual language (communication). Each of these measures uses an ordinal scale inclusive of no difficulty, some difficulty, a lot of difficulty, and cannot do at all.^33^

For this study, we use the five non-exclusive disability statuses common to both the ACS and WG-SS: hearing, vision, concentrating/remembering, mobility, selfcare. Each of these disability statuses was coded dichotomously (yes, no) for the ACS measures (corresponding to yes and no categories) and for the WG-SS measures per WG-SS recommendation^34^ and NHIS practice^27^ (yes = a lot of difficulty or cannot do at all; no = no difficulty or some difficulty). In addition, we created two distinct dichotomous composite measures, one for any ACS disability and one for any WG-SS disability, indicating whether the respondent had any of the five disability statuses (yes, no).

Two variables further specified the combinations of disability statuses reported with the ACS disability measures. The first indicated the ACS disability unique combinations reported using each of the five distinct ACS disability statuses (e.g., hearing, vision, & mobility; see Appendix A for all combinations). The second variable indicated the total number of reported ACS disability statuses reported: one, two, three or more.

One additional variable combined the ACS and WG-SS disability measures into a four-category nominal measure based on the ACS and WG-SS dichotomous composite measures: ACS disability only; WG-SS disability only; ACS and WG-SS disability; and no ACS or WG-SS disability.

#### Mortality

Mortality status is a dichotomous measure indicating whether the respondent was deceased through December 31, 2019.

#### Covariates

Demographic measures included age coded in single years, sex (female, male), and race-ethnicity (Non-Hispanic White, Non-Hispanic Black, Hispanic, Non-Hispanic American Indian/Alaskan Native, Non-Hispanic Asian/Pacific Islander, Non-Hispanic Other or Multiple races reported). Socioeconomic status measures included level of education (less than high school, high school/GED, some college or associates degree, bachelors degree or higher), poverty status (below, at/above poverty threshold), marital status (married/partnered, divorced/windowed/separated, never married), veteran status (yes, no). We also included measures for region (Northeast, North central, South, West), whether the respondent required assistance from a proxy to answer survey questions (yes, no), and year of survey (2011, 2012).

### Analytic strategy

Using the full analytic sample (N = 24,694), we first examined the percentage of respondents whose ACS disability status was documented in the WG-SS disability measures. To do so, we cross-tabulated the ACS individual, composite, and unique combinations disability statuses with the WG-SS individual and composite disability status measures. Preliminary examination of these results was used to determine whether to include particular disability status characteristics (e.g., number of disability statuses) in subsequent analysis.

We focused only on those respondents who reported an ACS disability (N = 4,821) to examine factors predicting documentation of ACS disability in WG-SS measures. Based on results from the preliminary analysis, we used a binary logistic model including a measure for number of ACS disability statuses, in addition to demographic characteristics, socioeconomic status, region, proxy status, and year of survey. We computed semi-partial correlations squared from this model to provide a standardized comparison of effects.

We then turned our attention to the consequences of the NHIS shifting from using the ACS disability measures to the WG-SS disability measures. To do so, we used the full analytic sample (N = 24,694) to estimate disability prevalence rates using: 1) the ACS disability status dichotomous composite measure; 2) the WG-SS disability status dichotomous composite measure; and 3) the ACS and WG-SS disability combined dichotomous measure. It is important to note that our estimates of prevalence are only inclusive of the five disability statuses examined in the study.

We used all cases with an eligible mortality status (N = 24,252) to examine a second consequence of shifting to use of the WG-SS disability measures, our understanding of comparative mortality risk. After converting the data to person years, we used Poisson regression models with a log link function of exposure time in study, calculated in months, to estimate predicted probabilities of mortality using: 1) the ACS disability status dichotomous composite measure; 2) the WG-SS disability status dichotomous composite measure; and 3) the ACS and WG-SS disability status four category composite measure.

All analysis was conducted with STATA 18.0 (College Station, TX). Computation of prevalence rates, as well as logistic and Poisson regression models adjusted for the complex sample design of the NHIS per National Center for Health Statistics (NCHS) recommendations. Examination of factors predicting whether ACS disability status was documented with the WG-SS disability measures using binary logistic models used unweighted data; computation of prevalence rates used sample adult weights; analysis of comparative mortality risk used sample adult mortality weights.

## Results

### Documentation of ACS disability in WG-SS measures

The percentage of ACS disability documented in WG-SS measures by ACS and WG-SS composite (any disability) and distinct (e.g., hearing, vision, etc.) disability status is reported in Table 1. Among respondents with any ACS disability status, 42.9% had their disability status documented in the WG-SS disability measures. There was some variation in the documentation of distinct ACS disability statuses in the WG-SS measures, with documentation highest for self-care disability (74.7%), lower for respondents with a vision (51.3%), concentrating/remembering (49.8%), or mobility (55.6%) disability, and lowest for those with a hearing disability (41.5%). Though not the focus of this study, as a point of comparison we note that 87.0% of respondents with a WG-SS disability status were documented as such in the ACS disability measures.

**Exhibit 1:** Percentage of ACS disability accurately documented in WG-SS measures by ACS and WG-SS composite and distinct disability status, 2011-2012 NHIS (N = 24,694)

| ACS disability status |  | % WG-SS disability status |  |  |  |  |  |
| --- | --- | --- | --- | --- | --- | --- | --- |
|  | N | Any | Hearing | Vision | Concentrating<br>/Remembering | Mobility | Selfcare |
| None | 19,873 | 1.55 | 0.28 | 0.34 | 0.25 | 0.78 | 0.04 |
| Any | 4,821 | 42.90 | 8.17 | 6.47 | 8.48 | 30.51 | 5.23 |
| Hearing | 1,560 | 41.54 | 23.08 | 5.90 | 7.18 | 24.04 | 4.29 |
| Vision | 1,051 | 51.28 | 9.42 | 25.21 | 11.42 | 30.26 | 7.71 |
| Concentrating<br>/Remembering | 1,625 | 49.78 | 9.42 | 8.55 | 23.57 | 31.38 | 9.35 |
| Mobility | 2,872 | 55.64 | 7.28 | 6.34 | 8.88 | 49.03 | 8.15 |
| Selfcare | 807 | 74.72 | 10.16 | 11.15 | 17.72 | 65.68 | 28.00 |

Note: ACS and WG-SS distinct disability status categories are not exclusive.

The percentage of ACS disability documented in WG-SS measures by ACS unique combinations and WG-SS composite and distinct disability status is reported in Appendix A. Preliminary analysis of these results revealed that rates of documentation of ACS disability status were comparatively lower among respondents with one disability status (10.3% - 41.0%), and comparatively higher among respondents with two or more disability statuses (48.3% - 100%). The only outliers to this pattern were for two of the unique combination groups, those with a hearing and vision disability (31.9%, 69 cases) and those with a hearing, vision, and self-care disability (0%, 1 case). Having observed this pattern, we also computed the percentage of documentation of ACS disability status by number of disability statuses (Appendix B). Among respondents with an ACS disability, documentation in the WG-SS measures was: 29.1% among the 2,884 respondents with one ACS disability status, 55.0% among the 1,130 respondents with 2 or more ACS disability statuses, and 75.2% among the 807 respondents with three or more ACS disability statuses.

Based on results of this preliminary analysis, we included the measure for number of ACS disability statuses in the logistic regression model examining factors predicting the documentation of ACS disability in WG-SS measures (Table 2). The distribution of all covariates by ACS disability status overall, and grouped by WG-SS status, are provided in Appendix C. Documentation of ACS disability in WG-SS measures was more likely for respondents with two, or three or more ACS disability statuses compared to those with one ACS disability status. Other covariates associated with a higher likelihood of documentation were age, female, living in the South compared to Northeast, and use of proxy respondent. Respondents with a bachelor’s degree or higher were less likely to have documentation than those with less than a high school degree. Based on analysis of semi-partial correlations squared, the strongest predictors of documentation of ACS disability in WG-SS measures were having two, or three or more ACS disability statuses.

**Exhibit 2:** Factors predicting the accurate documentation of ACS disability in WG-SS measures, 2011-2012 NHIS (N = 4,821)

|  | OR | 95% CI | Semi-partial correlations<sup>2</sup> |
| --- | --- | --- | --- |
| ACS number of disability statuses (ref = 1) |  |  |  |
| 2 | 2.73*** | (2.39, 3.12) | .037 |
| 3 or more | 6.19*** | (5.19, 7.38) | .082 |
| Age | 1.01*** | (1.01, 1.02) | .007 |
| Female (ref = male) | 1.31*** | (1.13, 1.52) | .002 |
| Race-ethnicity* (ref = White) |  |  |  |
| Black | 0.93 | (0.78, 1.11) | .000 |
| Hispanic | 1.00 | (0.80, 1.26) | .000 |
| American Indian/Alaskan Native | 0.71 | (0.43, 1.16) | .000 |
| Asian/Pacific Islander | 1.22 | (0.85, 1.75) | .000 |
| Other/Multiple Race | 1.40 | (0.90, 2.17) | .000 |
| Education (ref = <High school) |  |  |  |
| High school/GED | 0.97 | (0.83, 1.15) | .000 |
| Some college/Associates degree | 0.87 | (0.72, 1.04) | .000 |
| Bachelor degree or above | 0.74* | (0.59, 0.93) | .001 |
| Poverty (ref = at/above poverty threshold) |  |  |  |
| Below poverty threshold | 0.85* | (0.73, 1.00) | .002 |
| Marital status (ref = Married/Partnered) |  |  |  |
| Divorced/Widowed/Separated | 1.06 | (0.93, 1.22) | .000 |
| Single/Non Partnered | 1.06 | (0.88, 1.29) | .000 |
| Veteran (ref = nonveteran) |  |  |  |
| Veteran | 0.89 | (0.74, 1.08) | .000 |
| US Born (ref = yes) |  |  |  |
| No | 0.91 | (0.73, 1.14) | .000 |
| Region (ref = Northeast) |  |  |  |
| North Central | 1.06 | (0.83, 1.34) | .000 |
| South | 1.24* | (1.01, 1.53) | .001 |
| West | 1.25 | (0.99, 1.57) | .001 |
| Proxy (ref = Proxy not required) |  |  |  |
| Proxy required | 2.67*** | (1.95, 3.66) | .007 |
| Year (ref = 2011) | 0.97 | (0.85, 1.11) | .000 |
| Intercept | 0.17*** | (0.12, 0.25) |  |

Note: \* All racial-ethnic categories other than Hispanic are Non-Hispanic; ref = reference category.

### Consequence of using WG disability measures

#### Disability prevalence

Comparative prevalence of disability by ACS and WG-SS disability status are plotted in Figure 1. The prevalence of disability using the ACS composite measure was 17.0% (95% CI 16.3, 17.7). The prevalence of disability when using the WG-SS composite measure was 2.1 times lower at 8.1% (95% CI 7.7, 8.6). The prevalence of disability among respondents who reported a disability in either the ACS or WG-SS questions was 18.2% (17.5, 18.9).

**Exhibit 3:**
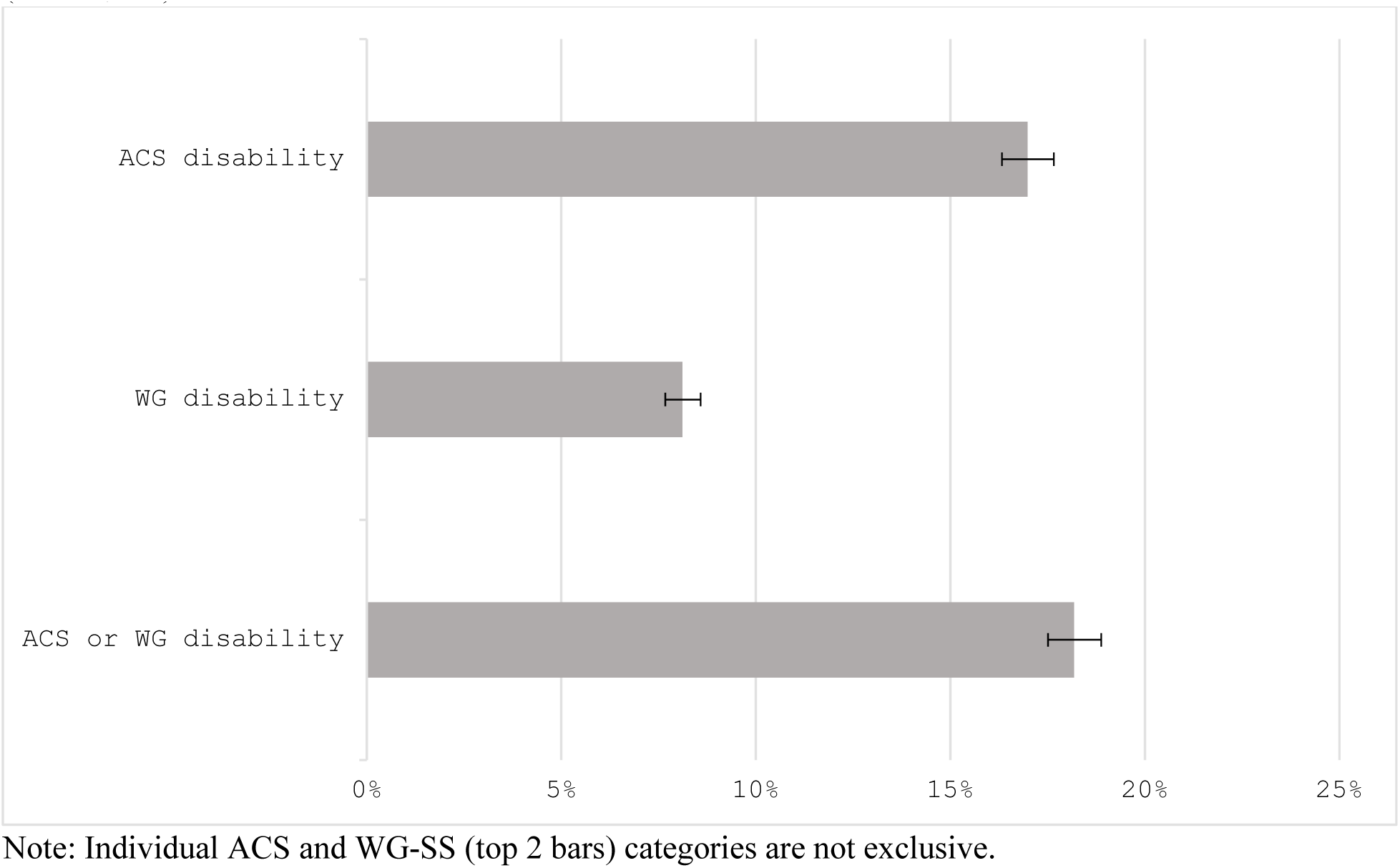
Comparative prevalence of disability by ACS and WG-SS disability status, 2011-2012 NHIS (N = 24,694)

#### Mortality risk

The predicted probabilities of mortality by ACS and WG-SS disability status are plotted in Figure 2. Results from the Poisson regression models used to compute the probabilities are provided in Appendix D. Most noticeably, when examining Exhibit 4, the predicted probability of mortality among disabled people was 1.3 times higher when using the WG-SS disability composite measure (Predicted probability (PP) = .020; 95% CI .018, .022) as opposed to the ACS disability composite measure (PP = .015; 95% CI .014, .016), with non-overlapping confidence intervals. To better understand why the WG-SS measure was associated with a comparatively higher mortality risk for disabled people, we also computed the predicted probability of mortality for each of the four categories of the ACS by WG-SS disability status measure. The predicted probability of mortality was equivalent among respondents whose ACS disability was not documented in the WG-SS measures (PP = .011, 95% CI .010, .013), and who reported only a WG-SS disability (PP = .012, 95% CI .008, .016). In contrast, it was 1.8 times higher among those respondents whose ACS disability was documented in the WG-SS measures (PP = .021, 95% CI .019, .023).

**Exhibit 4:**
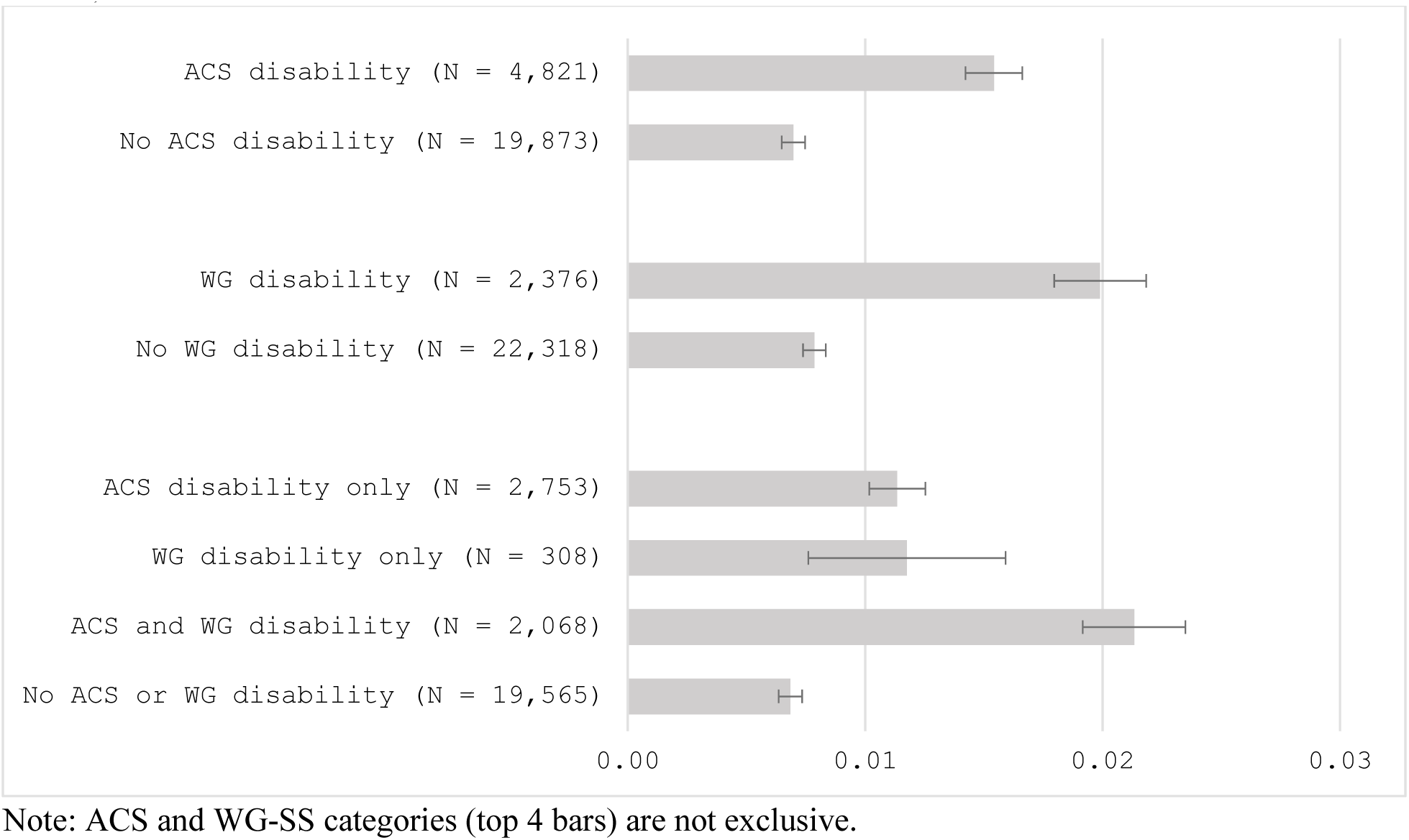
Predicted probabilities of mortality by ACS and WG disability status, 2011-2012 NHIS (N = 24,252)

## Discussion

Our aim in this paper was to evaluate the ramifications of switching from the ACS to the WG-SS disability questions in the NHIS. We did so with concern that this change may keep the NHIS from fulfilling its stated objective – to monitor the health of the population – for disabled people. Results from the study confirmed those reported in prior studies^26,28^ – over half of respondents who indicated they were disabled in the ACS questions were not documented as disabled in the WG-SS questions. Providing novel insight into which disabled adults are/are not being documented with the WG-SS questions, our analysis revealed that the WG-SS questions are more likely to document ACS disabled respondents as disabled if they had more than one disability status.

The second original contribution was to demonstrate that the change from using the ACS disability to the WG-SS disability questions had a direct impact on our understanding of this population. These results provide the first comparison of nationally representative estimates of non-institutionalized adult disabled population in the US from years when the ACS and WG-SS questions were fielded to the same NHIS samples. Use of the WG-SS questions to document disability status resulted in an underestimation of disability prevalence. Specifically, we found that the WG-SS questions report a disability prevalence that is 2.1 times lower than reported in the ACS questions.

Furthermore, we provide the first evidence that this underestimation of disabled people in the WG-SS questions has a direct impact on our understanding of a key population health outcome, mortality risk. Compared to the ACS questions, use of the WG-SS questions resulted in an overestimate of the comparatively higher mortality risk associated with disability status. It is likely that part of this mortality risk overestimation is due to the fact that the WG-SS questions are primarily documenting those with multiple disability statuses, which may be associated with higher mortality risk. In contrast, the ACS questions include the majority of those documented in the WG-SS questions, in addition to many disabled people with only one disability status – individuals who may have lower mortality risk.

These results contribute to growing literature examining the ACS and WG-SS disability questions. It is important to note that there is evidence that neither the ACS or WG-SS disability questions are fully sufficient in documenting disability status. Hall et al.^35^ recently reported that in the National Survey of Health and Disability (NSHD), a national sample of US adults with disabilities age 18 years and older that uses multiple measures to document disability status, including open-ended self-identification questions, both the ACS and WG-SS questions failed to capture 20% and 43% of people with disabilities, respectively. Specifically, both the ACS and WG-SS do not account for disabled people with intellectual and developmental disability, mental health disability, and chronic illness.^35^ The work by Hall et al. builds off of prior research documenting the limitations of the ACS and WG-SS disability questions,^23,28,36–38^ which highlight a long-term goal to expand and improve the questions used to assess disability.

However, a critical question remains. Until we have robust disability status questions that adequately document this population, which of the currently in-use disability questions, the ACS or WG-SS, does a better job of documenting more people with disabilities? Finding the answer to this question is an important next step in developing an improved disability questionnaire. Understanding whether the ACS or WG-SS better captures people with disabilities informs which of these questionnaires can be the ‘base’ on which additional questions can be added to capture more people with disabilities. Our comparison of the performance of these questions found that the ACS identified more people than the WG-SS, and a broader array of disabled people – specifically performing better in documenting disabled people with only one disability status – with disabilities in a nationally representative survey of community-dwelling American adults.

Results from this study demonstrate the impact of using the WG-SS in the NHIS on our understanding of the health of the disabled population in the US. Undercounting disabled people, as is the case when using the WG-SS questions in the NHIS, has critical impacts on multiple aspects of public health and social policy. People with disabilities face a multitude of health and economic inequities and disparities, making accurate estimates of disability prevalence for identifying and ensuring appropriate allocation of resources critical. We also found that compared to the ACS questions, the WG-SS questions overestimated mortality risk among people with disabilities, which certainly will require further investigations. The impact of inaccurate mortality rates cannot be understated. Not only does use of the WG-SS lead to an inaccurate understanding of comparative mortality risk among people with disabilities, we are concerned that an inflated mortality risk, as is the case when using the WG-SS questions, will do no more than affirm the false equivalence of disability with poor health among the medical community.^33^

While this study is among the first to directly compare the ACS to the WG-SS, there are limitations to consider when interpreting these findings. First, as noted above, there is evidence that both the ACS and WG-SS underestimate people with certain types of disabilities. Despite this limitation, our results are informative by identifying the ACS as a potentially more ideal option for adding questions that capture people with five specific disability statuses. Second, mortality was only assessed through 2019 which reflects the latest NHIS mortality data linkage available. While overall mortality rates would likely increase with longer follow up, we do not anticipate the direction of the association would change. These analyses are comparing the probability or mortality between people with disabilities the ACS and WG-SS and models were adjusted for factors that are associated with differences in disability prevalence and are increased mortality (age, sex, race-ethnicity). Third, the ACS and WG-SS were only fielded to the same subset of sample adults during two years, 2011 and 2012, which necessitated limiting the majority of our analysis to composite disability status. While it would be informative to extend this analysis to unique disability combinations, this was not possible with the size of the analytic sample.

## Conclusion

Our direct comparison of the ACS and WG-SS disability questionnaires in the NHIS suggests that the ACS identifies more people with disabilities, specifically performing better than the WG-SS in capturing those with one disability status. The disability prevalence determined using the WG-SS was 2.1 times lower than when using the ACS; the WG-SS also overestimated mortality risk among people with disabilities compared to the ACS. Based on these results, we are concerned that continued use of the WG-SS questions to document disability status imperils the ability of the NHIS to monitor the health of the disabled population in the US. While both the ACS and WG-SS undercount people with certain types of disabilities, our results suggest that among disability questions currently in use in us national surveys, the ACS would be a more ideal ‘base’ to add on questions that better capture a broader group of disabled people. These results have implications for policy and research, especially in health care and public health settings, and underscore an urgent need for investment in development of improved and expanded disability questionnaires.

## Data Availability

All data are available online at: https://nhis.ipums.org/nhis/

**Appendix A:**
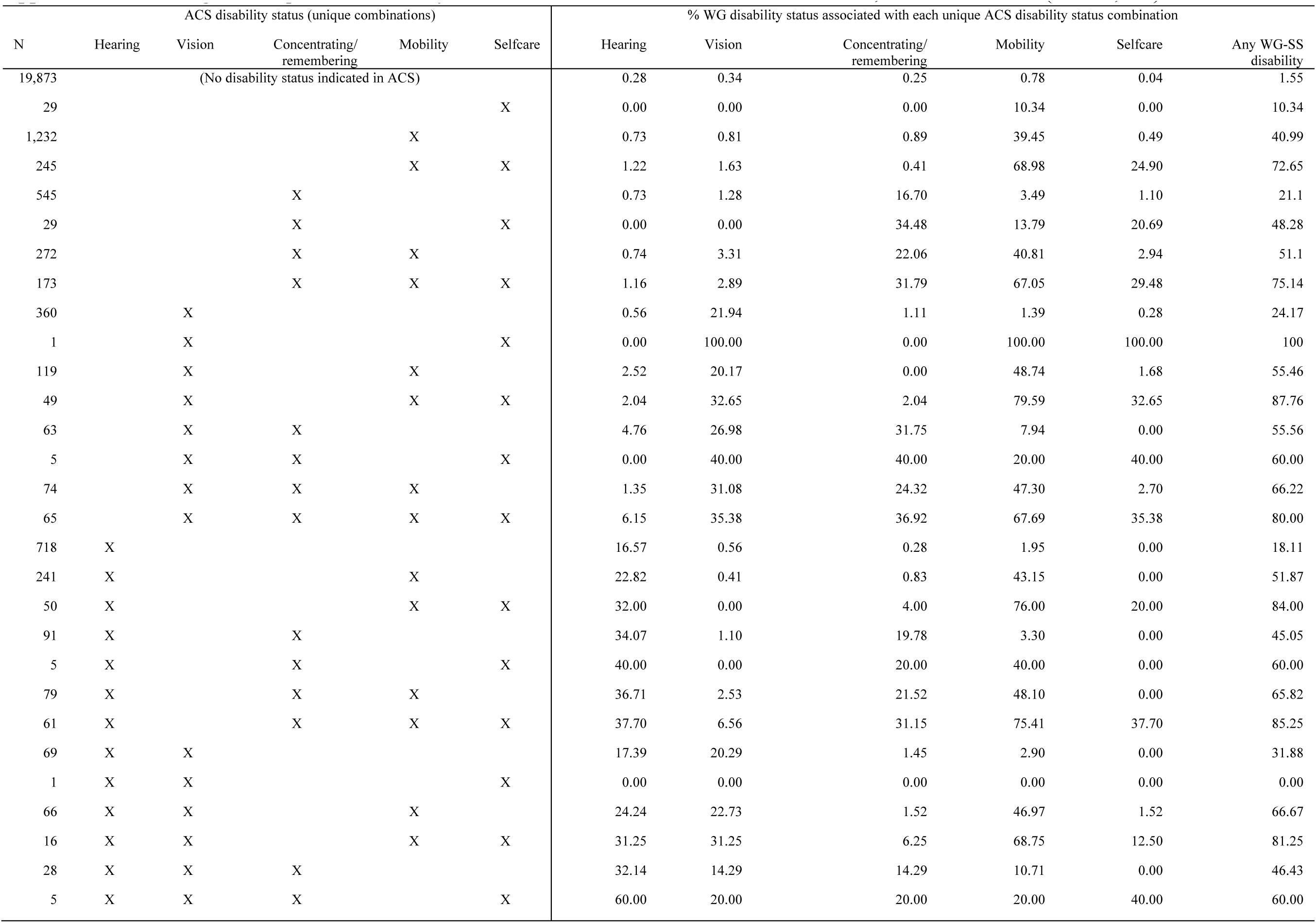
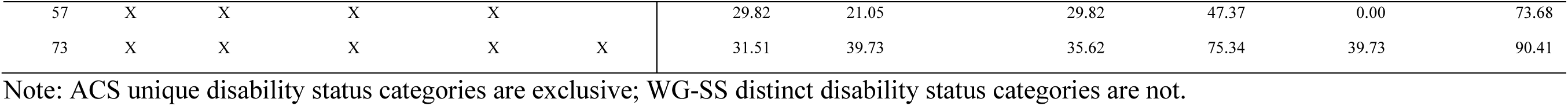
Percentage of unique ACS disability combinations documented in WG-SS measures, 2011-2012 NHIS (N = 24,694)

**Appendix B:** Percentage of ACS disability documented in WG-SS measures by number of disability statuses, 2011-2012 NHIS (N = 24,694)

| Number of ACS<br>disability statuses | N | WG-SS any<br>disability (%) |
| --- | --- | --- |
| 0 | 19,873 | 1.5 |
| 1 | 2,884 | 29.13 |
| 2 | 1,130 | 54.96 |
| 3 or more | 807 | 75.22 |

**Appendix C:** Distribution of all covariates by ACS disability status overall and grouped by WG-SS status, 2011-2012 NHIS (N = 4,930)

|  | All ACS<br>(N=4,821)<br>% | ACS only<br>(N=2,753)<br>% | ACS and WG-SS<br>(N=2,068)<br>% |
| --- | --- | --- | --- |
| Number of ACS disability statuses |  |  |  |
| 1 | 59.82 | 74.25 | 40.62 |
| 2 | 23.44 | 18.49 | 30.03 |
| 3 or more | 16.74 | 7.26 | 29.35 |
| Age (M) | 60.54 | 58.41 | 63.39 |
| Sex |  |  |  |
| Female | 56.15 | 52.74 | 60.69 |
| Male | 43.85 | 47.26 | 39.31 |
| Race-ethnicity |  |  |  |
| NHW | 62.83 | 64.15 | 61.07 |
| NHB | 17.47 | 17.04 | 18.04 |
| Hispanic | 13.79 | 13.44 | 14.26 |
| NHAIAN | 1.04 | 1.09 | 0.97 |
| NHAPI | 2.99 | 2.58 | 3.53 |
| NHO/M | 1.89 | 1.71 | 2.13 |
| Education |  |  |  |
| <HS | 27.80 | 24.63 | 32.01 |
| HS/GED | 29.58 | 28.88 | 30.51 |
| SC/AA | 28.36 | 30.04 | 26.11 |
| BA+ | 14.27 | 16.45 | 11.36 |
| Poverty threshold |  |  |  |
| Below | 28.00 | 25.57 | 31.24 |
| At/above | 72.00 | 74.43 | 68.76 |
| Marital status |  |  |  |
| Married/partnered | 38.50 | 41.01 | 35.15 |
| DWS | 45.01 | 41.34 | 49.90 |
| NM | 16.49 | 17.65 | 14.94 |
| Veteran status |  |  |  |
| Veteran | 15.93 | 16.67 | 14.94 |
| Nonveteran | 84.07 | 83.33 | 85.06 |
| US Born |  |  |  |
| Yes | 86.06 | 86.31 | 85.74 |
| No | 13.94 | 13.69 | 14.26 |
| Region |  |  |  |
| Northeast | 16.35 | 17.29 | 15.09 |
| North central/Midwest | 21.20 | 22.27 | 19.78 |
| South | 39.06 | 37.70 | 40.86 |
| West | 23.40 | 22.74 | 24.27 |
| Proxy status |  |  |  |
| Yes | 5.08 | 2.36 | 8.70 |
| No | 94.92 | 97.64 | 91.30 |
| Survey year |  |  |  |
| 2011 | 65.53 | 65.02 | 66.20 |
| 2012 | 34.47 | 34.98 | 33.80 |

**Appendix D:** Coefficients from Poisson regression models of mortality risk, 2011-2012 NHIS (N = 24,252)

|  | ACS only |  | WG-SS only |  | ACS-WG-SS categories |  | ACS or WG-SS |  |
| --- | --- | --- | --- | --- | --- | --- | --- | --- |
|  | b | 95% CI | b | 95% CI | b | 95% CI | b | 95% CI |
| ACS disability<br>(ref = no ACS disability) | 0.79*** | (0.69,<br>0.89) |  |  |  |  |  |  |
| WG disability<br>(ref = no WG disability) |  |  | 0.93*** | (0.82,<br>1.04) |  |  |  |  |
| ACS-WG categories<br>(ref = no ACS or WG disability) |  |  |  |  |  |  |  |  |
| ACS only |  |  |  |  | 0.51*** | (0.38,<br>0.63) |  |  |
| ACS & WG |  |  |  |  | 1.14*** | (1.01,<br>1.26) |  |  |
| WG only |  |  |  |  | 0.54** | (0.18,<br>0.90) |  |  |
| ACS or WG disability<br>(ref = no ACS or WG disability) |  |  |  |  |  |  | 0.80*** | (0.70,<br>0.90) |
| Intercept | -9.67*** | (-10.02,<br>-9.33) | -9.73*** | (-10.07,<br>-9.40) | -9.62*** | (-9.96,<br>-9.28) | -9.68*** | (-10.02,<br>-9.34) |

Note: Models adjusted for age, sex, race-ethnicity, and year.

## Notes

Potential conflicts of interest: Landes, Vaitsiakhovich – none; Swenor - CDC Health Equity Working Group Member

Funding: none

### Competing Interest Statement

Potential conflicts of interest: Landes, Vaitsiakhovich, none; Swenor, CDC Health Equity Working Group Member

### Funding Statement

This study did not receive any funding.

### Author Declarations

The study used ONLY openly available and de-identified human data that are located at: https://nhis.ipums.org/nhis/

